# Rapid, simplified whole blood-based multiparameter assay to quantify and phenotype SARS-CoV-2 specific T cells

**DOI:** 10.1101/2020.10.30.20223099

**Authors:** Catherine Riou, Georgia Schäfer, Elsa du Bruyn, Rene T. Goliath, Cari Stek, Huihui Mou, Deli Hung, Katalin A. Wilkinson, Robert J. Wilkinson

## Abstract

Rapid tests to evaluate SARS-CoV-2-specific T cell responses are urgently needed to decipher protective immunity and aid monitoring vaccine-induced immunity. Using a rapid whole blood assay requiring minimal amount of blood, we measured qualitatively and quantitatively SARS-CoV-2-specific CD4 T cell responses in 31 healthcare workers, using flow cytometry. 100% of COVID-19 convalescent participants displayed a detectable SARS-CoV-2-specific CD4 T cell response. SARS-CoV-2-responding cells were also detected in 40.9% of participants with no COVID-19-associated symptoms or who tested PCR negative. Phenotypic assessment indicated that, in COVID-19 convalescent participants, SARS-CoV-2 CD4 responses displayed an early differentiated memory phenotype with limited capacity to produce IFNγ. Conversely, in participants with no reported symptoms, SARS-CoV-2 CD4 responses were enriched in late differentiated cells, co-expressing IFNγ and TNFα and also Granzyme B. This proof of concept study presents a scalable alternative to PBMC-based assays to enumerate and phenotype SARS-CoV-2-responding T cells, thus representing a practical tool to monitor adaptive immunity in vaccine trials.

**Summary:** In this proof of concept study, we show that SARS-CoV-2 T cell responses are easily detectable using a rapid whole blood assay requiring minimal blood volume. Such assay could represent a suitable tool to monitor adaptive immunity in vaccine trials.

## INTRODUCTION

The outbreak of SARS-CoV-2 infection (causing the disease known as COVID-19) that first emerged in Wuhan, China in December 2019, was declared a global pandemic on 12^th^ March 2020, and is affecting all countries of the world, including those of Africa (Margolin et al., 2020). There is an urgent need to understand better the clinical manifestations and the pathogenesis of SARS-CoV-2 in order to develop relevant tools, including diagnostic tests, treatments and vaccines, to stop the spread of disease as well as strategies to best manage this disease in all population groups.

The clinical spectrum of COVID-19 is very wide, from asymptomatic through mild flu-like symptoms to severe pneumonia and death. Understanding what constitutes immune protection against SARS-CoV-2 is key to predicting long-term immunity and to inform vaccine design. While much emphasis has been placed on the B cell and antibody response, it is not yet clear what type of immune response confers protection to SARS-CoV-2 (Cox and Brokstad, 2020). Several studies suggest that the T cell response may play an important role in SARS-CoV-2 pathogenesis and reports indicating that patients lacking B cells can recover from SARS-CoV-2 infection further highlight the likely importance of T cell immunity (Altmann and Boyton, 2020); (Quinti et al., 2020). Additionally, accumulating evidence indicates that the presence of pre-existing, cross-reactive memory T cells specific for common cold coronaviruses may affect susceptibility to SARS-CoV-2 infection and partially explain the markedly divergent clinical manifestations of COVID-19 (Braun et al., 2020; Canete and Vinuesa, 2020; Lipsitch et al., 2020; Sette and Crotty, 2020).

Efficient, sensitive and simple assessment of human T-cell immunity remains a challenge. Most commonly used T cell assays necessitate the isolation of peripheral blood mononuclear cells (PBMC), requiring significant amount of blood. Therefore, whole blood assays could be more advantageous than PBMC-based methods, by significantly reducing blood volume (∼ 1 ml), making them more applicable to paediatric populations. Moreover, such assays are rapid, as they don’t require cell separation, and preserve the physiologic cellular and soluble environments, mimicking better human blood condition. Here we report a rapid (∼ 7 hrs) whole blood-based detection method of SARS-CoV-2-specific T cell responses with simple steps that could be adapted to settings of limited resources. This rapidly applicable assay could represent an easily standardizable tool to assess SARS-CoV-2-specific adaptive immunity to monitor T cell responses in vaccine trials, gain insight into what constitutes a protective response, or define the prevalence of SARS-CoV-2 T cell responders population wide.

## METHODS

### Study population

The population studied consisted of healthcare workers (HCW, n=31, 29% male) recruited between July and September 2020, from Groot Schuur Hospital in Cape Town, the hardest hit region of the initial COVID-19 epidemic in South Africa (Mendelson et al., 2020). Participants were classified according to i) reporting of COVID-19-associated symptoms, ii) whether a SARS-CoV-2 RNA PCR from a nasal or pharyngeal swab was performed and iii) SARS-CoV-2 PCR results. Based on these criteria, participants were subdivided into three groups: 1) persons with no COVID-19-associated symptoms (n=15); 2) persons who reported symptoms but tested SARS-CoV-2 PCR negative (n = 7); and 3) persons who had COVID-19-associated symptoms and tested SARS-CoV-2 PCR positive (n = 9) (**Table 1**). In all groups, the exposure to COVID-19 patients was comparable (86.8 to 100%). All participants with PCR confirmed COVID-19 had mild symptoms and did not require hospitalization. Blood samples were obtained a median of 7.3 weeks post-SARS-CoV-2 testing in persons with a negative test results and 4.7 weeks in those with a confirmed positive result (p=0.09). All participants were symptom-free at the time of sampling. In four SARS-CoV-2 PCR positive participants, a second sample was obtained 1 month later. The University of Cape Town’s Faculty of Health Sciences Human Research Ethics Committee approved the study (HREC: 207/2020) and written informed consent was obtained from all participants.

**Table 1:** Clinical characteristics of study participants.

|  | Healthcare Workers (n=31) |  |  |  |
| --- | --- | --- | --- | --- |
| n | 15 | 7 | 9 | P-value |
| Self-reported symptoms | No | Yes | Yes | - |
| Tested for COVID | No | Yes | Yes | - |
| Positive SARS-CoV-2 PCR | na | No | Yes | - |
| Positive SARS-CoV-2 N-spe IgG (%)<sup>a</sup> | 0% | 0% | 88.9% (8/9) | - |
| Presence of SARS nAbs (%)<sup>b</sup> | 0% | 0% | 100% | - |
| Contact with COVID patients (%) | 86.6% | 100% | 88.9% | ns |
| Time post PCR test (weeks)<sup>c</sup> | na | 7.3 [6.3-8.4] | 4.7 [2.6-7.2] | 0.09 |

<sup>a</sup> Measured using the Elecsys® Roche system <sup>b</sup> Measured using a SARS-CoV-2 pseudovirus neutralisation assay <sup>c</sup> Median and Interquartile range (IQR)

### Measurement of SARS-CoV-2 nucleocapsid-specific IgG in plasma

The measurement of SARS-CoV-2 specific antibodies was performed using the Roche Elecsys^®^ Anti-SARS-CoV-2 immunoassay (Roche Diagnostics, Basel, Switzerland). This semi-quantitative **e**lectro-chemiluminescent immunoassay measures SARS-CoV-2 nucleocapsid-specific IgG. The assay was performed by the South African National Health Laboratory Service (NHLS) and interpreted according to manufacturers’ instructions (Roche: V 1.0 2020-05). Results are reported as numeric values in form of a cut-off index (COI; signal sample/cut-off), where a COI < 1.0 corresponds to non-reactive plasma and COI ≥ 1.0 to reactive plasma. At 14 days post-SARS-CoV-2 PCR confirmation, the sensitivity and specificity of the Elecsys^®^ Anti-SARS-CoV-2 immunoassay is reported as 99.5% (95% CI, 97.0 to 100.0%) and 99.80% (95% CI, 99.69 to 99.88%), respectively (Favresse et al., 2020; Muench et al., 2020; National, 2020).

### SARS-CoV-2 pseudovirus neutralisation assay

Patient plasma was evaluated for SARS-CoV-2 neutralisation activity using a SARS-CoV-2 pseudovirus infection assay. Single-cycle infectious SARS-CoV-2 pseudovirions based on the HIV backbone expressing the SARS-CoV-2 Spike (S) protein and a firefly luciferase reporter were produced in HEK-293TT cells (Buck et al., 2004; Buck et al., 2005) by co-transfection of plasmids pNL4-3.Luc.R-.E- (NIH AIDS Reagent Program (#3418), Germantown, MD, USA) and pcDNA3.3-SARS-CoV-2-spike Δ18 (Rogers et al., 2020). HEK-293TT cell culture supernatants containing the virions were harvested 3 days post transfection and incubated with heat-inactivated patient plasma at 5-fold serial dilutions for 60 min at 37°C. Plasma/pseudovirus mixtures were then used for transfection of HEK-293T cells stably expressing the ACE2 receptor (Mou et al., 2020). Cells were lysed 3 days post infection using the Promega cell culture lysis reagent (Promega Biosciences Inc., San Luis Obispo, CA, USA) and assessed for luciferase activity using a GloMax^®^ Explorer Multimode Microplate Reader (Promega Biosciences) together with the Luciferase assay system (Promega Biosciences).

### Whole blood-based T cell detection assay

Blood was collected in sodium heparin tubes and processed within 3 h of collection. The whole blood-based SARS-CoV-2-specific T cell detection assay was adapted from a previously reported whole blood intracellular cytokine detection assay designed to quantitate *Mycobacterium tuberculosis* specific T cells in small volumes of blood (Hanekom et al., 2004). However, significant modifications have been made, including a reduced incubation time, the usage of a fixation buffer allowing the simultaneous lysis of red blood cells to streamline processing time, leading to faster acquisition of results (Riou et al., 2020). Here, we adapted this assay to detect SARS-CoV-2 specific T cells using synthetic SARS-CoV-2 PepTivator peptides (Miltenyi Biotec, Surrey, UK), consisting of 15-mer sequences with 11 amino acid overlap covering the immunodominant parts of the spike (S) protein, and the complete sequence of the nucleocapsid (N) and membrane (M) proteins. All peptides were combined in a single pool and used at a final concentration of 1 µg/ml. The workflow of the assay is presented in **Figure 1**. Briefly, 400 µl whole blood was stimulated with the SARS-CoV-2 S, N and M protein peptide pool at 37°C for 5 hrs in the presence of the co-stimulatory antibodies against CD28 and CD49d (1µg/ml each; BD Biosciences, San Jose, CA, USA) and Brefeldin-A (10µg/ml, Sigma-Aldrich, St Louis, MO, USA). Unstimulated blood was incubated with co-stimulatory antibodies, Brefeldin-A and an equimolar amount of DMSO. Red blood cell lysis and white cell fixation was then performed as a single step using a Transcription Factor Fixation buffer (eBioscience, San Diego, CA, USA) for 20 minutes. At this stage cells were cryopreserved in freezing media (50% foetal bovine serum, 40% RPMI and 10% dimethyl sulfoxide) and stored in liquid nitrogen until batched analysis.

**Figure 1:**
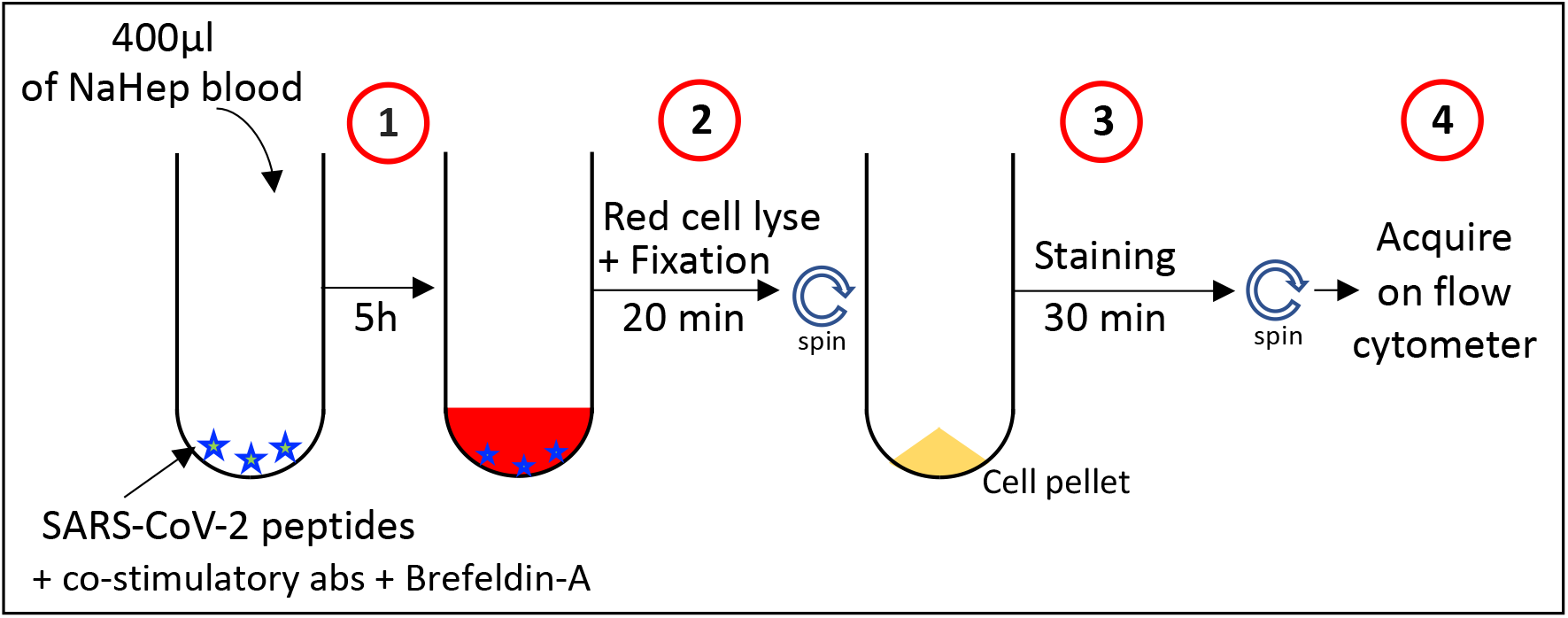
Schematic showing methodology and workflow of the whole blood assay for the detection of SARS-CoV-2-specific adaptive immune responses. **Step 1**: 400 µl of heparinized whole blood is incubated for 5 hours in the presence of a SARS-CoV-2-specific peptide pool in the presence of co-stimulatory antibodies (i.e. CD28 and CD49d) and Brefeldin-A. **Step 2**: Cells are incubated for 20 min in the presence of a transcription factor fixation buffer, leading to the simultaneous lysis of red blood cells and cell fixation. **Step 3:** Cells are stained for 30 min with an optimized panel of fluorophore labelled antibodies. **Step 4:** Samples are acquired on a flow cytometer. Control samples are processed with a similar workflow in the absence of SARS-CoV-2-specific peptide pool.

Cell staining was performed on cryopreserved cells that were thawed, washed and permeabilised with a Transcription Factor perm/wash buffer (eBioscience). Cells were then stained at room temperature for 30 min with antibodies for CD3 BV650, CD4 BV785, CD8 BV510, CD45RA Alexa 488, CD27 PE-Cy5, CD38 APC, HLA-DR BV605, Ki67 PerCP-cy5.5, PD-1 PE, Granzyme B (GrB) BV421, IFNγ BV711, TNFα PE-Cy7 and IL-2 PE/Dazzle 594, as detailed in **Supplementary Table 1**. Samples were acquired on a BD LSR-II and analysed using FlowJo (v9.9.6, FlowJo LCC, Ashland, OR, USA). A positive response was defined as any cytokine response that was at least twice the background of unstimulated cells. To define the phenotype of SARS-CoV-2-specific CD4 T cells, a cut-off of 30 events was used. The gating strategy is provided in **Supplementary Figure 1**.

### Statistical analyses

Graphical representations were performed in Prism (v8.4.3; GraphPad Software Inc, San Diego, CA, USA) and JMP (v14.0.0; SAS Institute, Cary, NC, USA). Statistical tests were performed in Prism. Non-parametric tests were used for all comparisons. The Kruskal-Wallis test with Dunn’s multiple comparison test was used for multiple comparisons and the Mann-Whitney and Wilcoxon matched pairs test for unmatched and paired samples, respectively.

## RESULTS

### Serological assessment of SARS-CoV-2 sensitization

Even though RT-PCR is the most specific technique to detect acute SARS-CoV-2 infection, the positivity rate drops rapidly as soon as 10 days post-symptom onset, particularly in individuals with mild forms of COVID-19 (Liu et al., 2020a; Liu et al., 2020b). Hence, serology assays provide an important complement to RNA testing to identify individuals who have been sensitized by SARS-CoV-2. Thus, to assess potential SARS-CoV-2 sensitization in participants who did not have a SARS-CoV-2 PCR test performed or tested PCR negative, the presence of SARS-CoV-2-specific antibodies (e.g. SARS-CoV-2 nucleocapsid-specific IgG) was measured and a SARS-CoV-2 pseudovirus neutralisation assay was also performed in all participants. While all participants with a positive SARS-CoV-2 PCR test exhibited *in vitro* anti-SARS-CoV-2 neutralizing activity and 8/9 (88.8%) had detectable SARS-CoV-2 nucleocapsid-specific IgG, none of the participants who tested negative or did not undergo PCR testing were positive for SARS-CoV-2 nucleocapsid-specific IgG (**Figure 2A**) or displayed robust *in vitro* anti-SARS-CoV-2 activity (**Figure 2B**). Overall, these results confirmed that PCR positive participants had been infected with SARS-CoV-2 and mounted an immune response to the virus and suggested that none of the participants who did not have a PCR test performed or tested PCR negative, despite experiencing COVID-19-like symptoms, have been SARS-CoV-2 infected.

**Figure 2:**
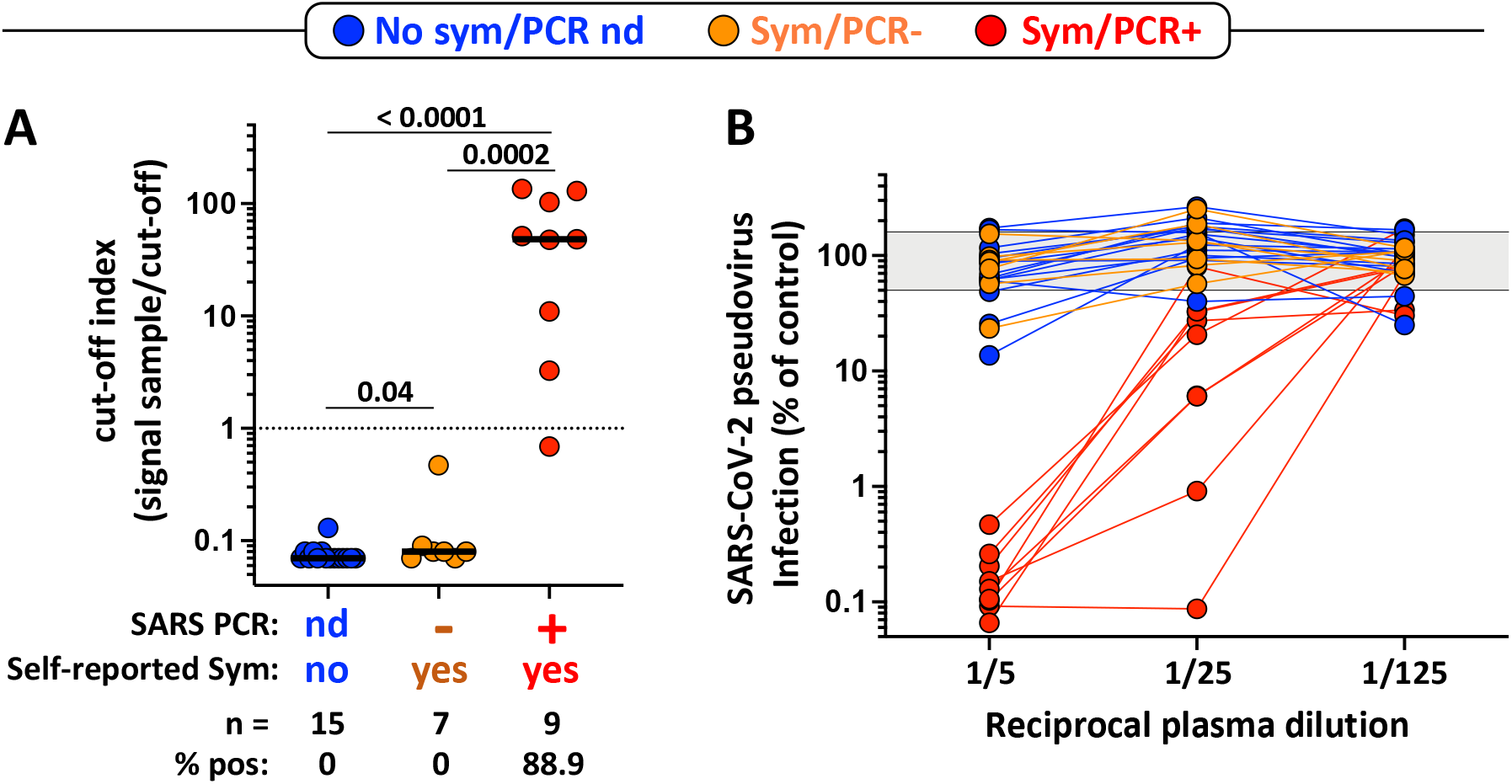
SARS-CoV-2 serological assessment. **A-** Quantification of SARS-CoV-2 Nucleocapsid-specific antibodies using the Elecsys^®^ Roche assay expressed as a cut-off index (signal sample/cut-off). Participants were grouped according to their clinical characteristics (Blue: no symptoms, no SARS-CoV-2 PCR performed; Orange: self-reported symptoms, SARS-CoV-2 PCR negative; and Red: self-reported symptoms, SARS-CoV-2 PCR positive). Bars represent the medians. The dotted line indicates the manufacturer’s cut-off value for positivity. Statistical comparisons were performed using a Mann-Whitney T-test. **B-** SARS-CoV-2 pseudovirus neutralization activity. SARS-CoV-2 pseudovirions pre-incubated with serially diluted patient plasma were used to infect ACE2-expressing HEK-293T cells. Luciferase activity as a measure for infection was assessed 3 days post-infection, and results are expressed as infection compared to control (untreated virions, grey shaded area) which was set 100%.

### Magnitude and functional profile of SARS-CoV-2-responding CD4 T cells

Amongst the 31 participants tested, 58% (n=18) had a detectable SARS-CoV-2-specific CD4 T cell response (producing any of the measured cytokines, IFNγ, TNFα or IL-2) using the described whole blood assay (**Figure 3A**). We then defined the magnitude and phenotype of SARS-CoV-2B-specific CD4 responses according to participants’ clinical characteristics (**Figure 3B&C**). All HCW with self-reported symptoms and a SARS-CoV-2 PCR positive test (n=9) were found to have SARS-CoV-2-specific CD4 T cells (median frequency: 0.47%, IQR: 0.28-0.65) (**Figure 3C**). Of note, the frequency of SARS-CoV-2 specific CD4 T cells did not associate with the magnitude of SARS-CoV-2 nucleocapsid-specific IgG (p = 0.64, r = −0.18) or plasma neutralizing activity (p = 0.46, r = 0.28) (data not shown). Interestingly, in participants with no detectable SARS-CoV-2 nucleocapsid-specific IgG or neutralizing antibody activity, of those with self-reported symptoms, 3/7 (43%) had detectable SARS-CoV-2-responding CD4 T cells (median of responders: 0.02%, IQR: 0.019-0.03) and 6/15 (40%) of those with no self-reported symptoms had detectable SARS-CoV-2-responding CD4 T cells (median of responders: 0.15%, IQR: 0.05-1.47), suggesting that these responses correspond to SARS-CoV-2 cross-reactive memory CD4 T cells.

**Figure 3:**
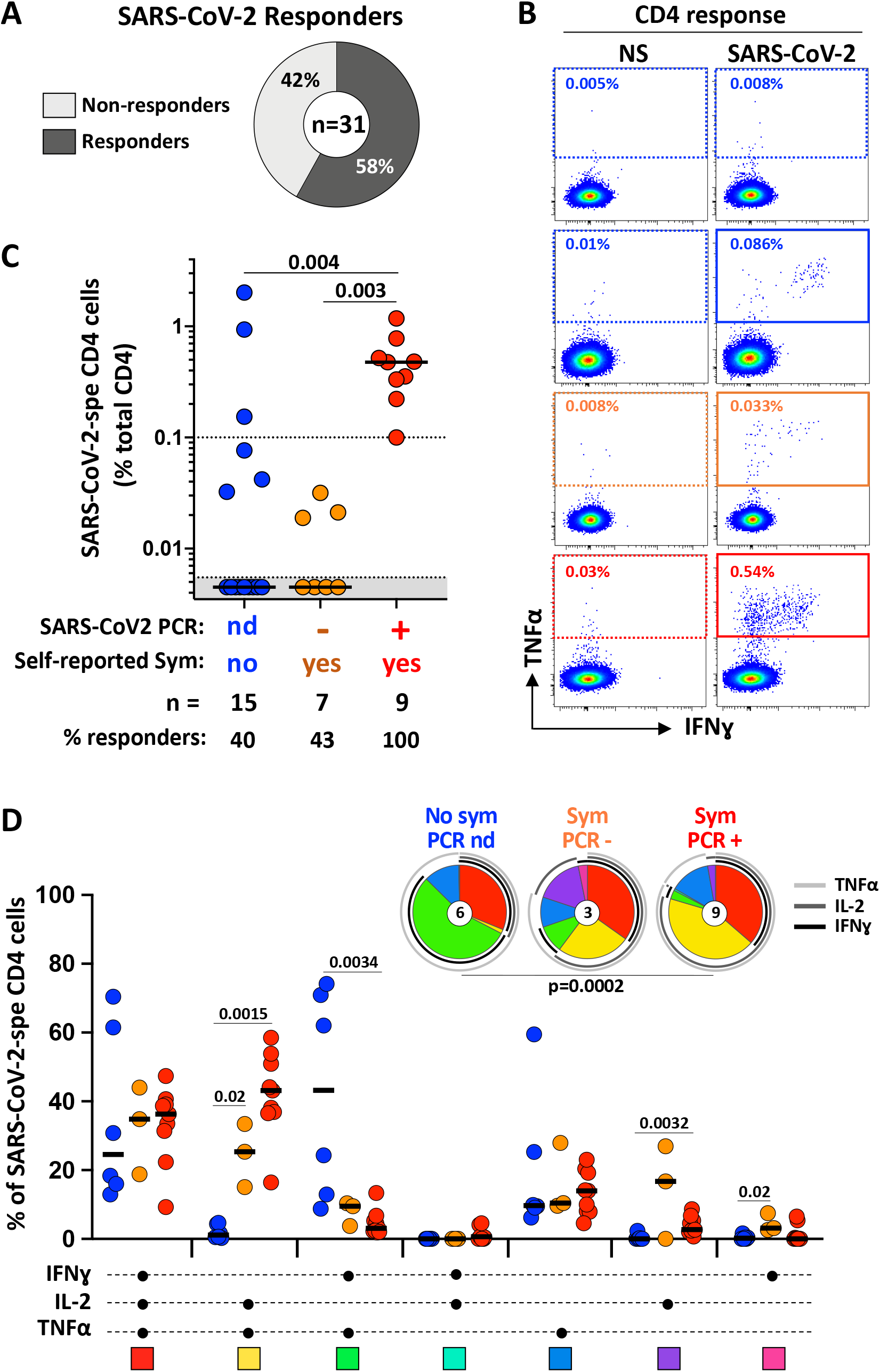
Magnitude and functional profile of SARS-CoV-2-specific CD4 T cells. **A-** Proportion of participants exhibiting a detectable SARS-CoV-2-specific CD4 T cell response. **B-** Representative examples of TNFα and IFNγ production in CD4 T cells in response to SARS-CoV-2 peptide pool. NS: no stimulation. **C-** Magnitude of SARS-CoV-2-specific CD4 T cell response (expressed as a percentage of total CD4 T cells) in participants grouped according to their clinical characteristics (Blue: no symptoms, no SARS-CoV-2 PCR performed; Orange: self-reported symptoms, SARS-CoV-2 PCR negative; and Red: self-reported symptoms, SARS-CoV-2 PCR positive). The number of participants and % of responders in each group is presented at the bottom of the graph. Statistical comparisons were performed using the Kruskal-Wallis test. **D-** Polyfunctional profile of SARS-CoV-2-specific CD4 T cells in each group. The x-axis displays the composition of each combination which is denoted with a dot for the presence of IL-2, IFNγ and TNFα. The medians (black bar) are shown. Each combination is color-coded, and data are summarized in the pie charts, where each pie slice represents the median contribution of each combination to the total SARS-CoV-2 response. The arcs identify the contribution of TNFα (light grey), IL-2 (grey) and IFNγ (black) to SARS-CoV-2 response. The Wilcoxon Rank Sum Test was used to compare response patterns between groups. Statistical differences between pie charts were defined using a permutation test.

Next, we defined the polyfunctional profile of SARS-CoV-2-responding CD4 T cells, based on their capacity to co-express IL-2, IFNγ or TNFα (**Figure 3D**). The overall functional profile of SARS-CoV-2-specific cells in PCR positive participants was distinct from participants with no reported symptoms (p=0.0002). Indeed, the SARS-CoV-2-specific CD4 response in PCR+ participants was characterized by limited expression of IFNγ and was enriched in cells co-expressing IL-2 and TNFα. On the contrary, in participants reporting no symptom, most SARS-responding CD4 cells were distributed between triple functional cells (IL-2+IFNγ+TNFα+) and cells co-producing IFNγ and TNFα. Overall, in this assay TNFα was the predominant cytokine produced, with its production being significantly higher compared to IL-2 (p = 0.0026) and IFNγ (p < 0.0001) (**Supplementary Figure 2**).

### Phenotypic assessment of SARS-CoV-2-responding CD4 T cells

While the proposed assay can be performed using a limited antibody panel to identify the frequency of SARS-CoV-2-responding T cells, by solely measuring cytokine production, the use of a more extensive antibody panel also permits definition of the phenotypic profile of these cells. To this end, we included additional markers to assess the memory differentiation (CD27, CD45RA), cytotoxic potential (GrB) and activation profile (HLA-DR, CD38, Ki67, PD-1) of SARS-CoV-2-responding CD4 T cells.

**Figure 4A&B** shows that in all individuals with symptoms, SARS-CoV-2-specific CD4 T cells displayed almost exclusively (median: 97.7%) an early differentiated memory phenotype (ED: CD45RA-CD27+). On the contrary, in 4/6 HCW with no symptoms, SARS-CoV-2-specific CD4 T cells exhibited a predominant late differentiated phenotype (LD: CD45RA-CD27-).

**Figure 4:**
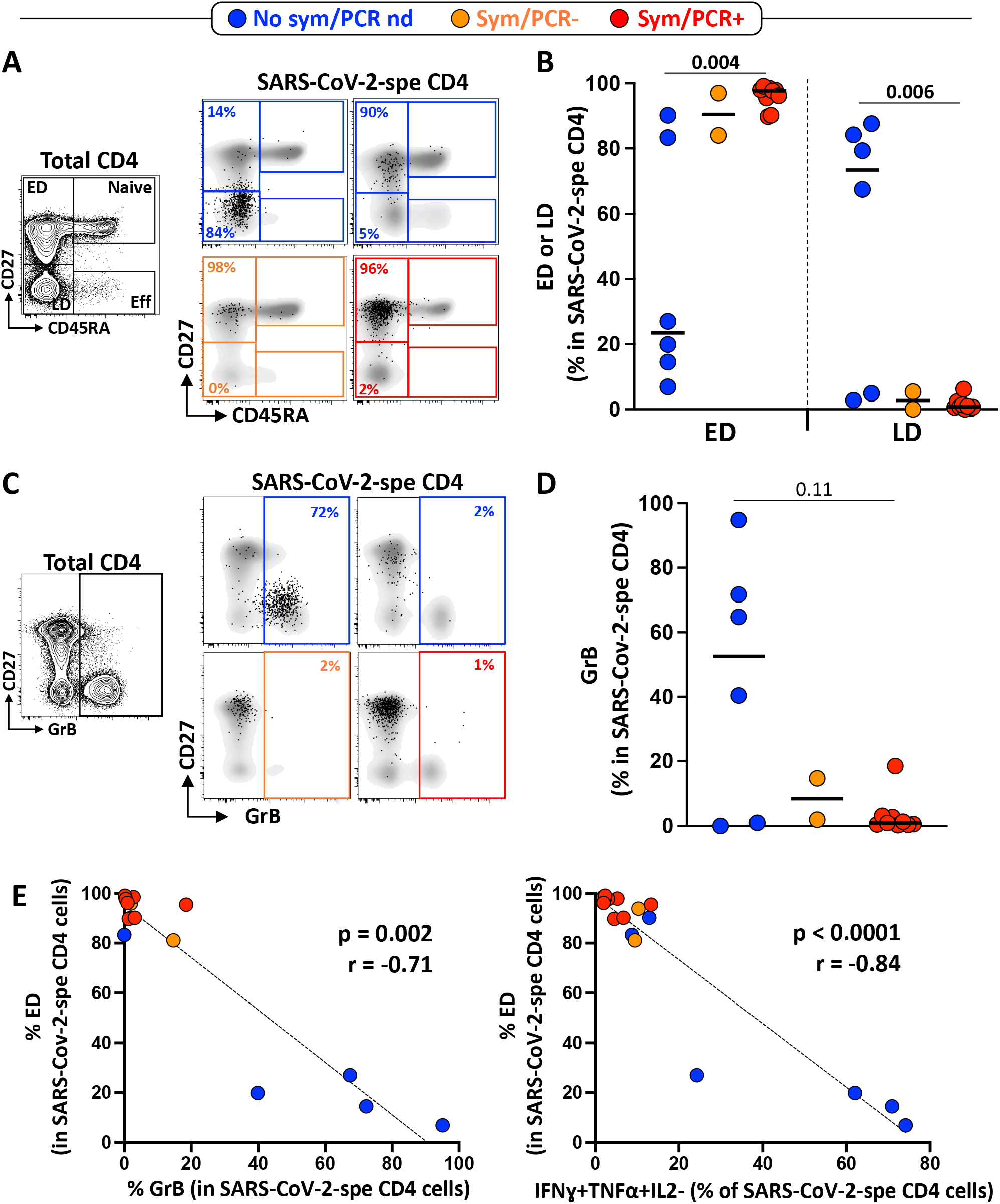
Memory differentiation profile and Granzyme B (GrB) expression in SARS-CoV-2-specific CD4 T cells. **A-** Representative examples of the memory differentiation profile of SARS-CoV-2-specific CD4 T cells based on the expression of CD45RA and CD27. The flow plot on the right shows the distribution of Naïve (CD45RA+CD27+), Early differentiated (ED: CD45RA-CD27+), Late differentiated (LD: CD45RA-CD27-) and Effector (Eff: CD45RA+CD27-) in total CD4 T cells. **B-** Summary graph of the proportion of ED and LD in SARS-CoV-2-specific CD4 T cells in each group (Blue: no symptoms, no SARS-CoV-2 PCR performed; Orange: self-reported symptoms, SARS-CoV-2 PCR negative and Red: self-reported symptoms, SARS-CoV-2 PCR positive). Statistical comparisons were performed using the Kruskal-Wallis test. **C-** Representative examples of GrB expression in total and SARS-CoV-2-B specific CD4 T cells. **D-** Summary graph of GrB expression in SARS-CoV-2-specific CD4 T cells in each group. Statistical comparisons were performed using the Kruskal-Wallis test. **E**- Relationship between the proportion of ED within SARS-CoV-2-specific CD4 T cells and GrB expression or the proportion of IFNγ+TNFα+IL2-SARS-CoV-2-specific CD4 T cells. Correlations were tested by a two-tailed non-parametric Spearman rank test.

While the role of cytotoxic CD4 T cells is still unclear, the presence of these cells has been described in several viral infections (Juno et al., 2017). We thus measured GrB expression in SARS-CoV-2-responding CD4 T cells. While GrB was barely detectable in SARS-CoV-2-specific CD4 T cells in PCR positive participants, elevated GrB expression was observed in 4 out of 6 participants without symptoms (**Figure 4C&D**). Moreover, the proportion of ED SARS-CoV-2-responding CD4 T cells inversely associated with GrB expression (p = 0.002, r = −0.71) and the proportion of IFNγ and TNFα dual producing cells (p < 0.0001, r = −0.84) (**Figure 4E**). The phenotypic characteristics of GrB expressing SARS-CoV-2-responding CD4 T cells (i.e. highly differentiated and producing IFNγ and TNFα) are in agreement with a recent report describing CMV-specific CD4 CTL T cells (Pachnio et al., 2016).

To determine if the overall phenotypic profile of SARS-CoV-2-responding CD4 T cells allowed discrimination of participants based on their clinical characteristics, we performed a hierarchical clustering analysis (**Figure 5A**) and a principal component analysis (PCA) (**Figure 5B**), including four parameters (e.g. proportion of IFNγ+TNFα+IL2+ and IFNγ-TNFα+IL2+ cells, proportion of ED, and GrB expression). Both analyses show that participants who reported no symptoms and were negative for SARS-CoV-2 Abs separated clearly from PCR positive participants. Conversely, PCR and SARS-CoV-2 antibody negative HCWs reporting symptoms could not be separated from PCR positive participants. The loading plot shows that GrB expression and the proportion of triple positive cells were the main drivers permitting the segregation of participants who reported no symptoms and were negative for SARS-CoV-2 Abs (**Figure 5C**)

**Figure 5:**
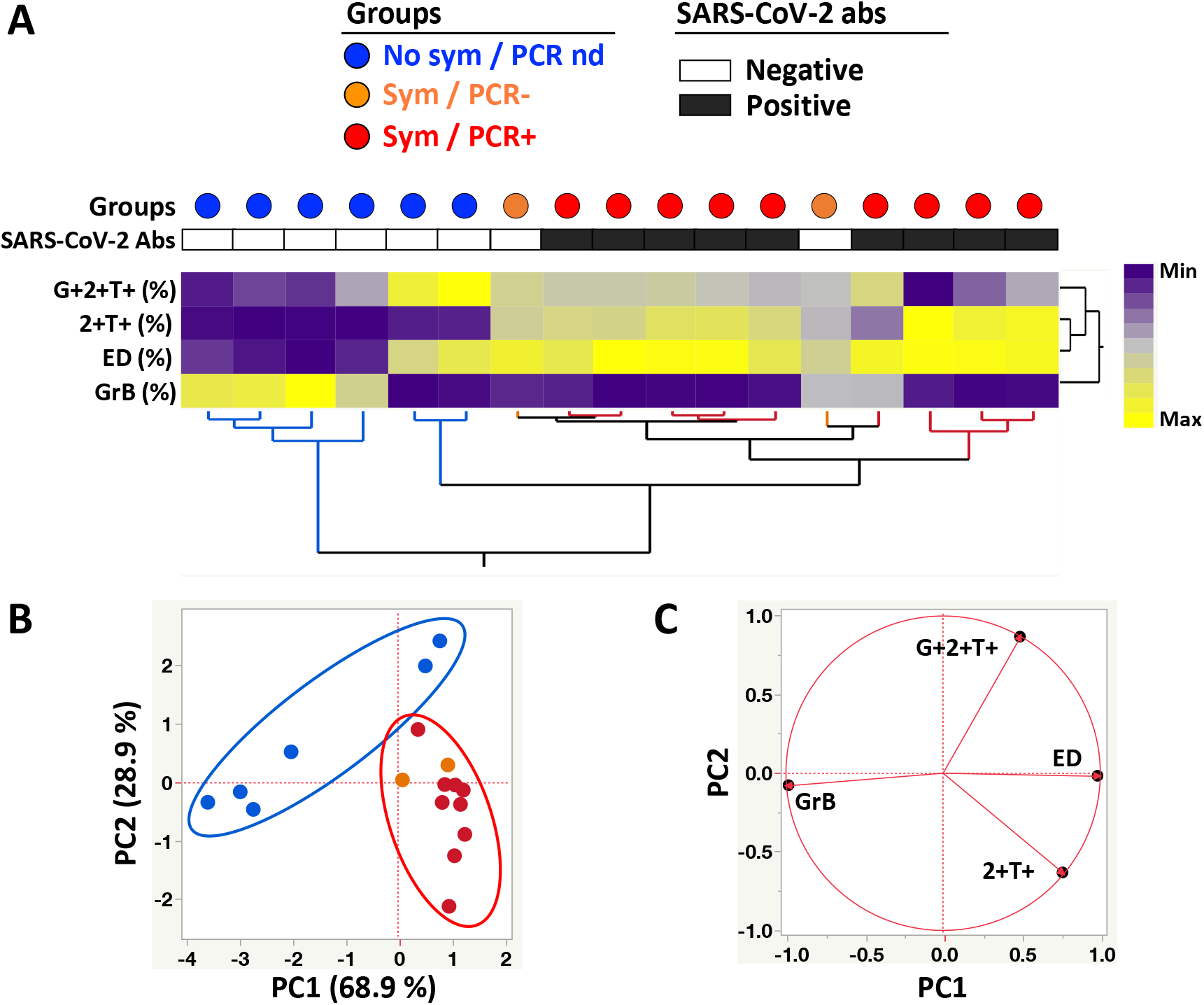
Phenotypic signature of SARS-CoV-2-specific IFNγ+ CD4+ T cells according to clinical characteristics. **A-** Non-supervised two-way hierarchical cluster analysis (HCA, Ward method) using three phenotypic parameters (i.e. GrB expression and the proportion of ED and IFNγ+TNFα+IL2-) from SARS-CoV-2-specific CD4 T cells. Each column represents a participant and is color-coded according to their clinical characteristics indicated by a dot at the top of the dendrogram (Blue: no symptoms, no SARS-CoV-2 PCR performed; Orange: self-reported symptoms, SARS-CoV-2 PCR negative; and Red: self-reported symptoms, SARS-CoV-2 PCR positive). Participants with a positive SARS-CoV-2 serology test are indicated by a black box and a white box identifies SARS-CoV-2 antibody-negative subjects. Data are depicted as a heatmap coloured from minimum to maximum values for each parameter. **B-** Principal component analysis on correlations, derived from the three studied parameters. Each dot represents a participant. The two axes represent principal components 1 (PC1) and 2 (PC2). Their contribution to the total data variance is shown as a percentage. **C-** Loading plot showing how each parameter influences PC1 and PC2 values.

Lastly, as the expression of activation markers (HLA-DR, CD38, PD-1 or Ki67) on antigen-specific T cells are indicative of active infection (Chen and Wherry, 2020; Mathew et al., 2020), we defined the expression of these markers on SARS-CoV-2-responding CD4 T cells. No significant difference was observed in any of the measured markers amongst the different groups, notwithstanding a few outliers observed in the PCR positive participant group (**Figure 6A**).

**Figure 6:**
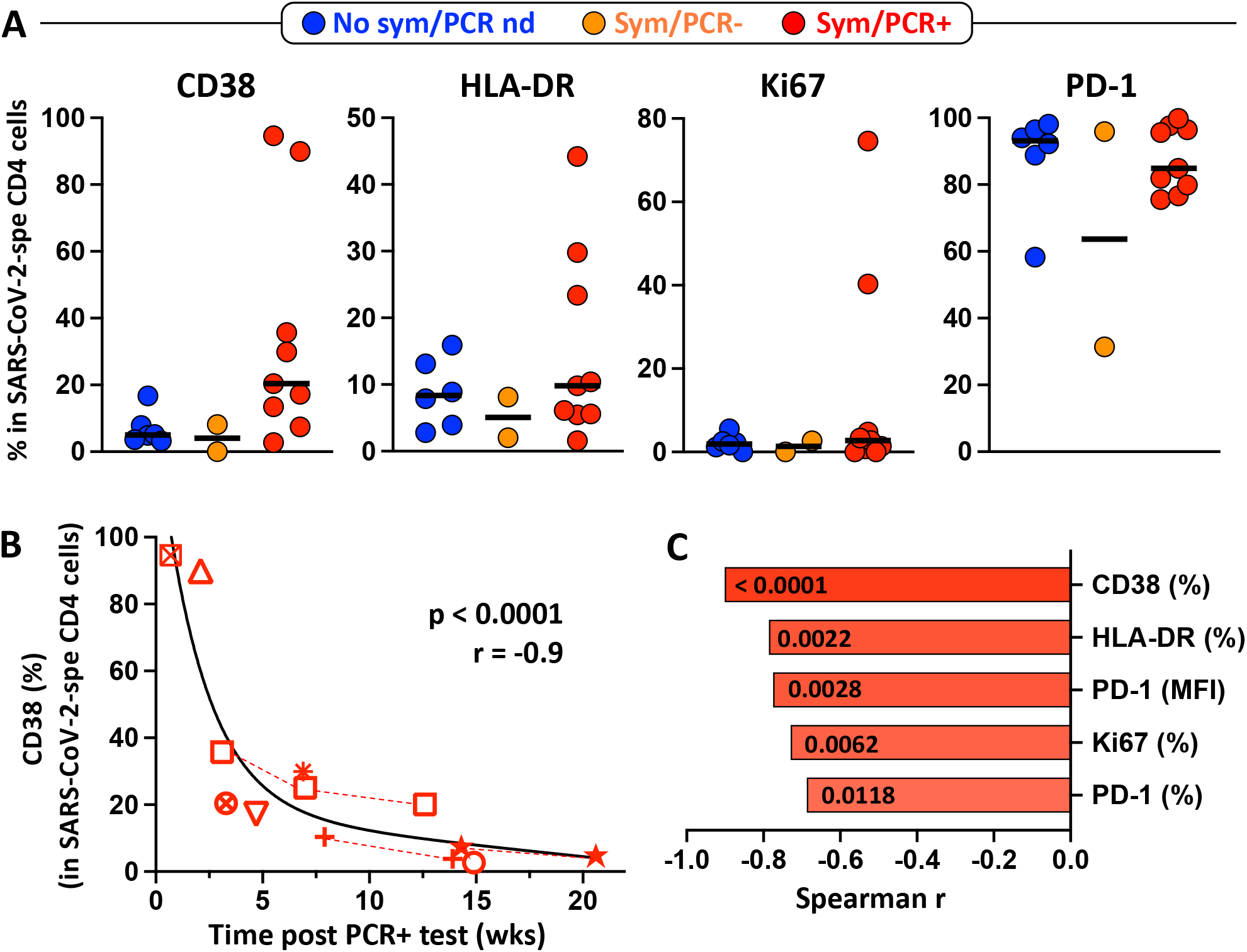
Activation profile of SARS-CoV-2-specific CD4 T cells. **A-** CD38, HLA-DR, Ki67 and PD-1 expression in SARS-CoV-2-specific CD4 T cells in each group (Blue: no symptoms, no SARS-CoV-2 PCR performed; Orange: self-reported symptoms, SARS-CoV-2 PCR negative and Red: self-reported symptoms, SARS-CoV-2 PCR positive). No significant differences were observed between groups for any markers, using the Kruskal-Wallis test. **B-** Association between CD38 expression in SARS-CoV-2-specific CD4 T cells and the time post SARS-CoV-2 PCR positive test (weeks). Each symbol represents a participant (n=9). Dotted red lines identify participant with longitudinal samples. Correlations were tested by a two-tailed non-parametric Spearman rank test. **C-** Comparison of the correlation between the time post SARS-CoV-2 PCR positive test (weeks) and the expression of different activation profile markers, ranked according to the strength of the association. Spearman correlation r values are plotted on the x-axis and corresponding *P-*values are shown within each bar.

Interestingly, regardless of the participants’ clinical characteristics, PD-1 expression was highly expressed on SARS-CoV-2-responding CD4 T cells, raising the question of a potential intrinsic dysfunction/exhaustion of coronavirus-specific memory CD4 T cells (Jubel et al., 2020; Wherry and Kurachi, 2015; Wykes and Lewin, 2018). We then defined the relationship between the activation profile of SARS-CoV-2-specific CD4 T cells and time post positive PCR test.

**Figure 6B** shows that in PCR positive participants, the expression of CD38 associated strongly with the time post PCR testing (p <0.0001, r = −0.91), where CD38 expression decreased sharply in the first 2 to 3 weeks post testing, suggesting a rapid clearance of the pathogen. Similar associations where observed for all tested activation markers, with CD38 expression showing the strongest correlation (**Figure 6C**).

## DISCUSSION

In this proof of concept analysis assessing the use of a simple whole blood assay to measure SARS-CoV-2-specific T cell responses in a small health care worker cohort, we show that SARS-CoV-2-specific CD4 T cells (expressing IFNγ, TNFα or IL-2) were easily detectable in all tested convalescent COVID-19 participants. However, in participants who did not experience any COVID-19-related symptoms or tested SARS-CoV-2 PCR negative (despite reporting symptoms), 9/22 (40.9%) also exhibited a detectable SARS-CoV-2-specific CD4 T cell response. Nevertheless, the median frequency of SARS-CoV-2-specific CD4 T cells in the latter groups was ∼ 5-fold lower compared to SARS-CoV-2 PCR positive participants. Importantly, unlike convalescent COVID-19 participants, none of participants without symptoms or who tested PCR negative had detectable SARS-CoV-2 nucleocapsid-specific IgG or displayed robust neutralizing activity. This strongly suggests that in these participants, SARS-CoV-2-responding CD4 T cells may correspond to a pre-existing cross-reactive memory CD4 T cell response. Thus, the sole presence SARS-CoV-2-reactive CD4 T cells does not permit to infer SARS-CoV-2 infection or sensitization. These findings are in line with a number of studies demonstrating the presence of SARS-CoV-2-reactive CD4 T cells in 40 to 60% of SARS-CoV-2-unexposed individuals in different populations around the world (Grifoni et al., 2020; Le Bert et al., 2020; Rydyznski Moderbacher et al., 2020; Sekine et al., 2020). Additionally, it is important to point out that all these studies were performed using PBMC and it is encouraging to find that a whole blood-based assay using limited amount of blood (< 1ml) yields comparable results.

Further analyses of the polyfunctional and phenotypical profile of SARS-CoV-2-specific CD4 T cells revealed that regardless of the clinical characteristics of the participants, the most prevalent cytokine detected in response to SARS-CoV-2 peptides was TNFα. These observations are in accordance with other studies pointing out an impairment of SARS-CoV-2 adaptive immune responses, characterized by low level of type I and type II interferon (Blanco-Melo et al., 2020; Sattler et al., 2020). However, it is also possible that the limited IFNγ production, we observed in this study, is related to the short stimulation time (e.g. 5hrs) used in the assay. Nevertheless, this suggests that TNFα could be a more reliable target than IFNγ, to detect SARS-CoV-2-specific CD4 T cell responses. Additionally, SARS-CoV-2-responding CD4 T cells in all participants were characterized by elevated of PD-1 expression. Such profiles have been observed in acute SARS-CoV-2 infection and is likely driven by ongoing viral replication (Schub et al., 2020; Sekine et al., 2020; Zheng et al., 2020). However, PD-1 expression on antigen-specific T cells has been shown to decrease upon pathogen clearance (Barber et al., 2006; Sester et al., 2008). Thus, elevated PD-1 expression in COVID-19 convalescent persons and uninfected responders could reflect an intrinsic state of functional exhaustion of SARS-CoV-2-reactive memory CD4 T cells.

Lastly, SARS-CoV-2-responding CD4 T cells were qualitatively different between convalescent COVID-19 participants and participants who did not experience COVID-19-associated symptoms. In the former group, SARS-CoV-2-specific CD4 T cells displayed almost exclusively an early differentiated memory phenotype; while in the latter group, SARS-CoV-2-responsive CD4 T cells exhibited a late differentiated memory phenotype and were enriched in GrB. Differences in the attributes of SARS-CoV-2-reactive T cells distinguishing recent SARS-CoV-2 infection from pre-existing immunity have been recently reported: strong *ex vivo* Elispot responses and enhanced proliferation capacity were frequently observed in confirmed COVID-19 patients but rare in pre-pandemic or unexposed seronegative persons (Ogbe et al., 2020). This is consistent with the phenotypic features we observed in our study. It remains to be determined whether the presence of pre-existing cross-reactive memory T cells offers any protection against SARS-CoV-2.

Although our results rely on a small number of participants and need to be confirmed in larger cohorts, this study suggests that a whole blood assay could represent a valuable tool to assess SARS-CoV-2-specific adaptive immunity. The whole blood assay described here is rapid and scalable for varying complexities depending on laboratory resources and questions to be addressed. It can be adapted to a simple 4-colour flow cytometry assay to monitor the frequency of T cell responses in epidemiological studies or vaccine trials. Alternatively, it can also be used to characterize in more depth the specific attributes of SARS-CoV-2-specific T cells, as shown here, to aid deciphering cellular features that may associate with immune correlates of protection. Finally, as this standardizable assay requires a limited volume of blood, it can be easily used in COVID-19 paediatric cases.

## Data Availability

all data is available upon request

## Acknowledgment

The authors thank all the participants.

## Author contributions

CR, KAW and RJW designed the study. CR performed the whole blood assay and flow cytometry experiments and analysed and interpreted the data. GS performed the neutralizing assay experiments. EdB, RTG and CS recruited the study participants. KAW, HM and EL provided critical reagents. CR and KAW wrote the manuscript with all authors contributing to providing critical feedback.

---

**Supp table 1:** Description of the antibody panel used in the study.

| Markers | Fluorochrome | Clone | Company | Cat. Number | Role |
| --- | --- | --- | --- | --- | --- |
| CD3 | BV650 | OKT3 | BioLegend | 317323 | Lineage |
| CD4 | BV785 | OKT4 | BioLegend | 317428 |  |
| CD8 | BV510 | RPA-T8 | BioLegend | 301048 |  |
| CD45RA | Alexa 488 | HI100 | BioLegend | 304114 | Memory differentiation |
| CD27 | PE-Cy5 | 1A4CD27 | Beckman | 6607107 |  |
| CD38 | APC | HIT2 | BD Bioscience | 555462 | Activation |
| HLA-DR | BV605 | L243 | BioLegend | 307640 |  |
| Ki67 | PerCP-Cy5.5 | B56 | BD Bioscience | 561284 |  |
| PD-1 | PE | EH12.2H7 | BioLegend | 329906 |  |
| GrB | BV421 | GB11 | BD Bioscience | 563388 | Cytotoxic potential |
| IFNγ | BV711 | 4S.B3 | BioLegend | 502540 | Functions |
| TNFα | PE-Cy7 | MAB11 | BioLegend | 502930 | |
| IL-2 | PE/Dazzle™ 594 | MQ1-17H12 | BioLegend | 500344 |  |

**Supp Figure 1:**
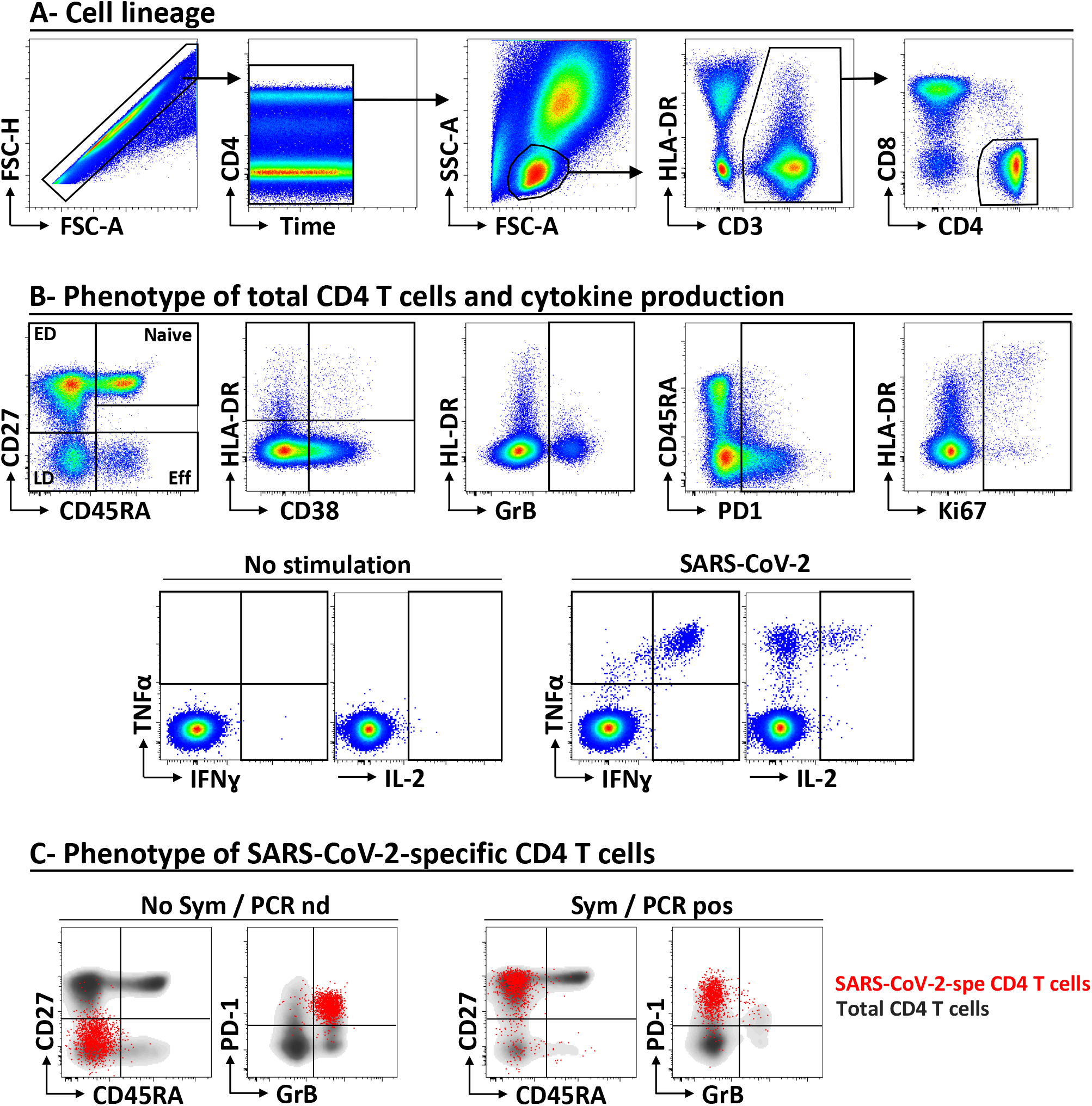
Gating strategy used to identify SARS-CoV-2 specific CD4 T cells and defined their phenotypic characteristics. **A-** Cell lineage. **B-** Phenotyping of total CD4 T cell and cytokine production in response to SARS-CoV-2 peptides. **C-** Phenotype of SARS-CoV-2 responding CD4 T cells in one participant with no symptom (left) and one COVID-19 confirmed participant (right). SARS-CoV-2-specific CD4 T cells (expressing any measured cytokine) are depicted in red.

**Supp Figure 2:**
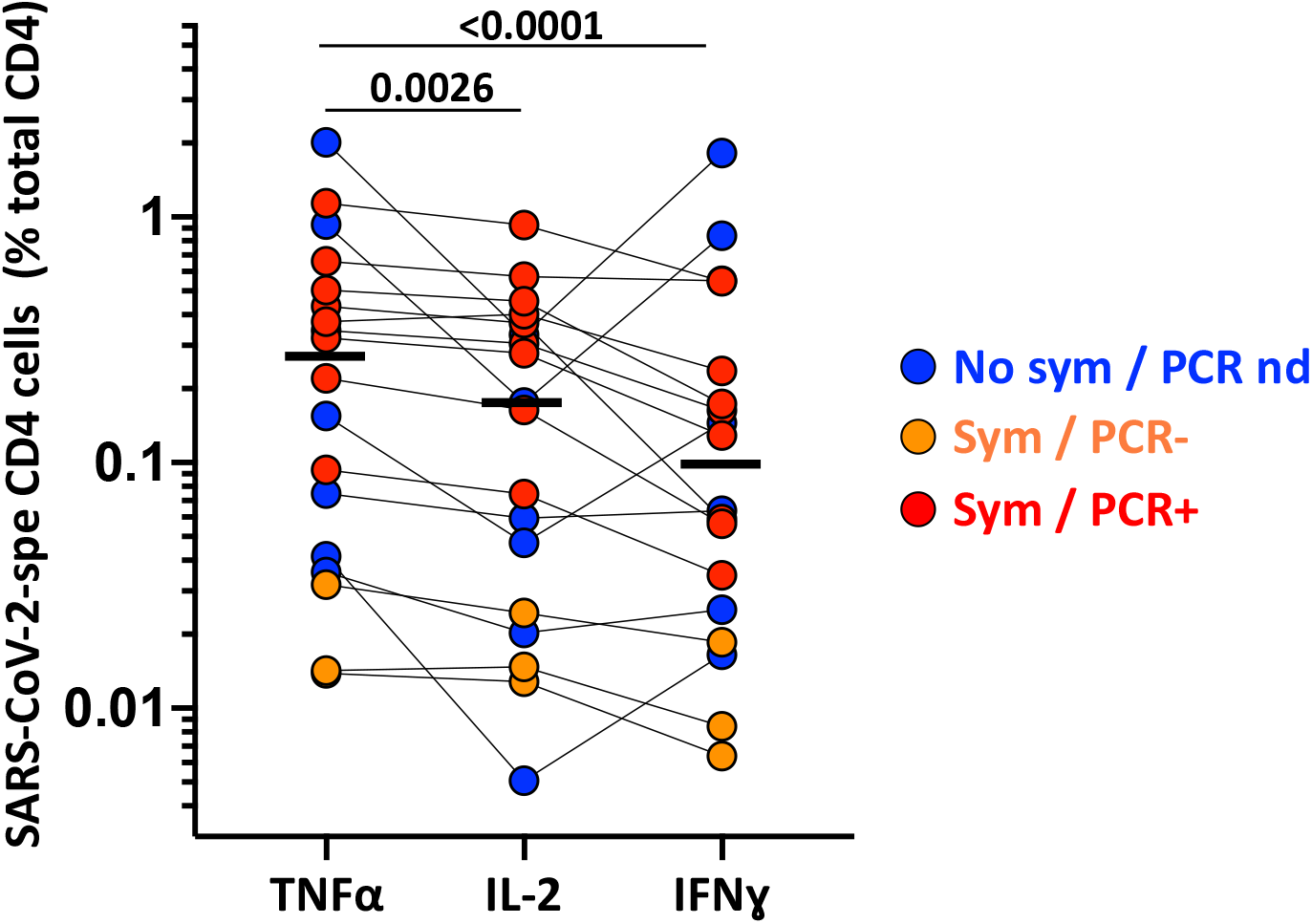
Comparison of magnitude of SARS-CoV-2-specific CD4 T cell response expressing TNFα, IL-2 or IFNγ in participants exhibiting a detectable response to SARS-CoV-2 peptide pool. Each dot represent a participant and is color-coded according to clinical characteristics (Blue represents no symptoms, no SARS-CoV-2 PCR performed; Orange: self-reported symptoms, SARS-CoV-2 PCR negative and Red: self-reported symptoms, SARS-CoV-2 PCR positive). Black bars indicate medians. Statistical comparisons were performed using a nonparametric paired Friedman test.

